# Retrospective Machine Learning Approach for Forecasting In-Hospital Death in ICU Patients After Cardiac Arrest

**DOI:** 10.1101/2025.05.05.25327009

**Authors:** Yong Si, Li Sun, Shuheng Chen, JunYi Fan, Elham Pishgar, Kamiar Alaei, Greg Placencia, Maryam Pishgar

**Author notes:** These authors contributed equally to this work (Yong Si, Li Sun). **Emails:** Yong Si. Greg Placencia: Emial.

## Abstract

Accurate identification of patients at high risk of in-hospital mortality in intensive care units (ICUs) is vital for enhancing clinical decision-making and improving patient care strategies. As traditional statistical models often fall short in modeling nonlinear and multifactorial clinical variables, this study explores a machine learning (ML) approach to overcome these limitations.

Our research performed a retrospective study using the MIMIC-IV database, focusing on 2,385 ICU patients who met predefined eligibility criteria. Numerical features were summarized through statistical aggregations (maximum, minimum, mean), while categorical attributes underwent structured encoding. The dataset was split into 70% for training and 30% for validation. We applied a combination of regularization techniques (LASSO, Ridge, ElasticNet) and Random Forest-based importance ranking for feature selection.

Multiple supervised ML algorithms, including CatBoost, XGBoost, and Support Vector Machines, were benchmarked using metrics such as AUC-ROC, calibration plots, and decision curve analysis. SHAP values were employed to enhance model explainability.

The CatBoost algorithm achieved the most favorable results with AUC scores of 0.904 and 0.868 on the training and test sets. These findings suggest that the proposed model offers a reliable, interpretable, and potentially integrable solution for ICU mortality risk prediction.

## 1 Introduction

Cardiac arrest (CA) continues to pose a major public health burden worldwide due to its substantial rates of mortality. In the US alone, more than 350,000 individuals suffer from out-of-hospital cardiac arrest (OHCA) each year, yet the survival rate to hospital discharge remains low at approximately 10.8%, with in-hospital survival reported at 26.4% [1, 2]. Additionally, more than 300,000 hospitalized patients suffer in-hospital cardiac arrest (IHCA) annually in the U.S., with a survival rate to discharge of approximately 25% [3]. IHCA incidence ranges from 9 to 10 cases per 1,000 hospital admissions, and its mortality rate is alarmingly high, between 80% and 100% [4]. Most deaths occur during acute events; however, some patients die after successful initial resuscitation due to post-cardiac arrest syndrome, characterized by neurological and multi-organ dysfunction. Consequently, current clinical guidelines underscore the critical importance of promptly identifying patients at elevated risk of experiencing cardiac arrest. [4–6].Recent works have similarly explored outcome prediction in critically ill populations with various complications such as stroke and sepsis using machine learning-based methods [7, 8].

Although predictive models exist, their accuracy often remains unsatisfactory due to limitations such as small sample sizes (¡1000) or the absence of C-statistic evaluation [9–11]. Current clinical guidelines recommend neuron-specific enolase (NSE) as the principal biomarker for neurological prognosis [12–17]. Moreover, ICU readmission prediction and stroke risk models built on MIMIC data have demonstrated the potential of structured ICU datasets for risk modeling [18, 19].Recent research has demonstrated that the electronic Cardiac Arrest Risk Triage (eCART) score is associated with a notable reduction in in-hospital cardiac arrest (IHCA) events; however, its reliance on laboratory test results poses challenges for routine clinical application. [20]. Similarly, traditional scoring systems like the Modified Early Warning Score (MEWS) encounter limitations due to short prediction windows and insufficient sensitivity [21].

Different from traditional prediction models that rely on pre-selected variables, Machine Learning (ML) methods can efficiently integrate numerous variables computationally, thereby enhancing predictive accuracy, and simultaneously employ multiple variable screening techniques to improve model precision and efficiency, enabling more timely risk prediction [22].The gradient boosting family, particularly CatBoost, XGBoost, and LightGBM, has shown strong potential in clinical prediction [23, 24]. compared LASSO regression, XGBoost, logistic regression, and NEWS 2 for predicting in-hospital mortality in ICU patients with cardiac arrest, finding ML models to outperform traditional scoring systems [25–27].

In recent research, ML techniques applied to ICU datasets, including MIMIC database, have yielded strong performance in predicting complications such as acute kidney injury and short-term survival, highlighting the practical relevance of ML-based approaches [28, 29].Additionally, MIMIC-IV, an open-access ICU database from Beth Israel Deaconess Medical Center (2008–2019), provides rich clinical data, including demographics, laboratory results, vital signs, medications, and outcomes, enabling large-scale retrospective analyses [30, 31].

This study develops an interpretable ML model for ICU cardiac arrest mortality prediction by integrating advanced feature selection and data augmentation. Our research employ LASSO, ridge regression, ElasticNet, and random forest-based ranking to identify key predictors. Numerical features are augmented via statistical aggregations to capture dynamic physiological changes. SHAP analysis enhances interpretability, fostering clinical adoption and improving patient outcomes.The main contributions of this study are as follows:

- We propose a comprehensive feature selection strategy combining LASSO, Ridge, ElasticNet, and Random Forest importance rankings to distill 17 clinically meaningful predictors from the full ICU stay data.
- Through benchmarking eight machine learning models with extensive hyperparameter tuning via GridSearchCV, we demonstrate that CatBoost achieves the best predictive performance (AUC = 0.904 training, 0.868 testing), surpassing traditional classifiers and validating model calibration and discrimination.
- We employ SHAP analysis to elucidate key predictive factors such as Glasgow Coma Scale (GCS), serum lactate, and arterial blood pressure minimum, thus enhancing model interpretability and supporting clinical adoption.

## 2 Methods

### 2.1 Data Extraction and Study Design

This retrospective study utilized data from the MIMIC-IV database, a publicly accessible and high-fidelity critical care resource comprising detailed clinical records from 2008 to 2019. The dataset was developed under the oversight of MIT, Cambridge, MA, USA and Beth Israel Deaconess Medical Center. Access to the database was granted upon successful completion of the National Institutes of Health, with data extraction and study conduct authorized under Certification Number 50778029. The use of a large, diverse critical care dataset enhances the generalizability of the model to varied clinical settings, addressing a major limitation of earlier small-cohort studies.

This study implemented a structured machine learning pipeline comprising cohort selection, data preprocessing, feature engineering, model development, and validation. Adult ICU patients were identified from the MIMIC-IV database based on cardiac arrest diagnosis and admission criteria. Numerical features were summarized using statistical aggregations (minimum, maximum, mean), and categorical variables were label-encoded. Missing values were imputed using a Random Forest-based approach to preserve multivariate dependencies and reduce bias.

To construct a robust and interpretable feature set, multiple selection techniques were applied, including LASSO, Ridge, and ElasticNet regularization, alongside tree-based importance ranking from Random Forests. The intersection of selected variables yielded a clinically relevant subset used to train and evaluate eight machine learning models: CatBoost, XGBoost, AdaBoost, Random Forest, Logistic Regression, SVM, Naïve Bayes, and Neural Network. Among them, CatBoost achieved the highest performance across AUC, precision, recall, and F1-score metrics.

A four-part evaluation framework was adopted to ensure clinical reliability: model calibration was assessed via Brier score and calibration plots; t-tests were used to detect distributional shifts between training and test sets; SHAP analysis enabled both global and local interpretability; and hyperparameter tuning was conducted through grid search with five-fold cross-validation. This framework supports scalable and interpretable prediction of in-hospital mortality, offering a practical tool for early identification of high-risk ICU patients. The overall workflow is illustrated in Figure 1.

**Figure 1.**
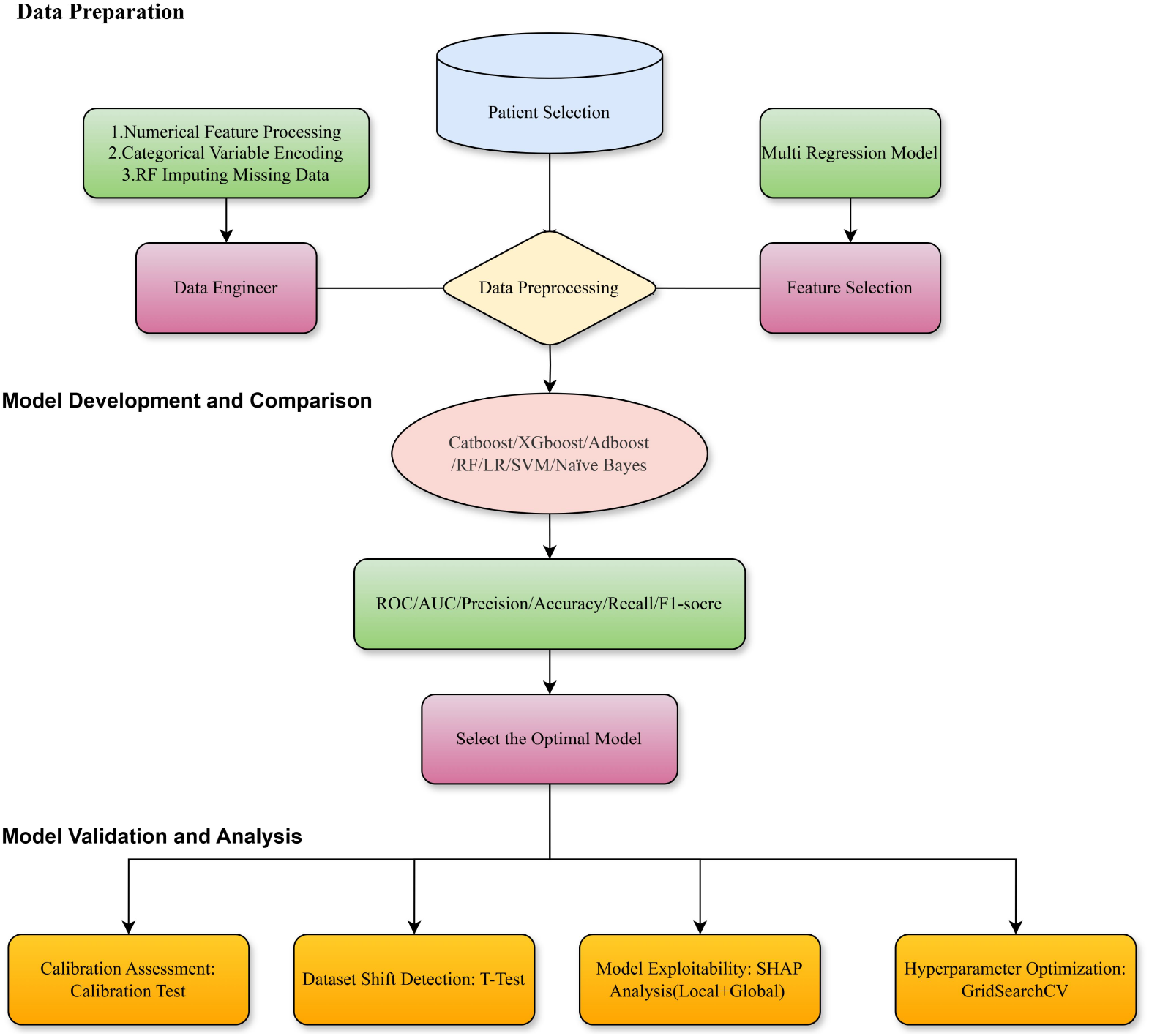
Flowchart of study design.

### 2.2 Patient Selection

Patients with a diagnosis of cardiac arrest (CA) were identified from the MIMIC-IV database using ICD-9 code 4275 and ICD-10 codes I46, I462, I468, and I469. Individuals aged 18 years or older at the time of ICU admission were eligible for inclusion, whereas patients lacking documented ICU admission records were excluded from the analysis. A total of 2,608 patients were initially identified, and after applying exclusion criteria, 2,385 adult patients were included in the final cohort. The study dataset was then randomly divided into 70% a training set (n = 1,670) and a 30% test set (n = 715). The patient selection process is illustrated in Algorithm 1.

#### Algorithm 1

**Patient Selection for ICU Mortality Prediction**

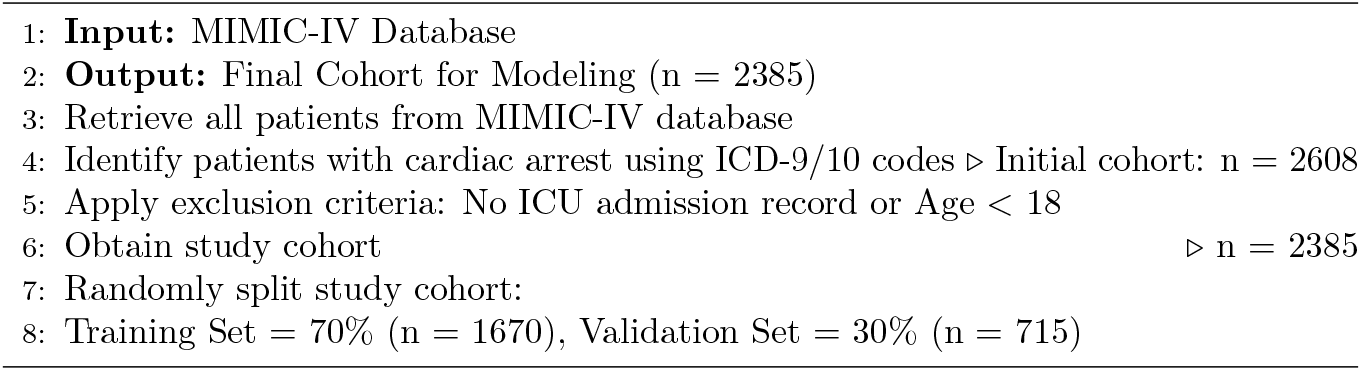

### 2.3 Feature Engineering

Demographics, vital signs, laboratory tests, medical history, interventions, and scoring system variables were extracted from the MIMIC-IV database using structured query language (SQL) with MySQL. The prediction model incorporated only clinical and laboratory variables recorded during the entire stay of ICU. Similar to approaches in other large-scale structured data domains, such as public transit networks, statistical summarization techniques were employed to reduce dimensionality while retaining meaningful dynamic features [32].For numerical variables, the maximum, minimum, and average values were extracted for further analysis. Categorical variables were processed as binary indicators, where the presence of a condition (e.g., heart failure) was assigned a value of 1, and its absence was assigned 0. Race was categorized into White, Black, Unknown, and Others. Temporal aggregation of numerical data (min, max, mean) captured ICU trajectory patterns, providing clinically meaningful dynamic predictors. Encoding comorbidities as binary indicators aligned with clinical practice for risk stratification. Choosing compact label encoding over one-hot encoding preserved dataset tractability for real-time clinical deployment.

Feature selection was based on prior research [33–35], clinical relevance, and data availability. Demographic characteristics, including age at hospital admission, sex, admission type, admission location, and insurance status, were selected because they represent baseline patient profiles and social determinants that significantly impact survival outcomes after cardiac arrest. Vital signs, such as systolic blood pressure (SBP), mean blood pressure (MBP), respiratory rate (RR), heart rate (HR), diastolic blood pressure (DBP), body temperature, and oxygen saturation (oxygen saturation (SpO_2_)), were incorporated as they provide real-time assessments of hemodynamic stability and respiratory function, critical indicators of early physiological deterioration. Comorbidities, including heart failure, arrhythmia, myocardial infarction, and pulmonary disease, were captured to account for chronic disease burden, which independently contributes to post-arrest mortality risk. Laboratory test results, including anion gap, hemoglobin, platelet count, sodium, potassium, lactate, INR, blood urea nitrogen (BUN), creatinine, chloride, glucose, prothrombin time (PT), and white blood cell count, were extracted to reflect the patient’s underlying metabolic, renal, hematologic, and systemic inflammatory status, which are key determinants of ICU prognosis. Medical interventions such as cardiopulmonary resuscitation (CPR), defibrillation, intubation, and ventilation were included to capture the severity of clinical events and the aggressiveness of life-sustaining treatments administered, both of which are strong prognostic indicators. Scoring systems including the Simplified Acute Physiology Score III (SAPS III), Sequential Organ Failure Assessment (SOFA) score, and the Glasgow Coma Scale (GCS) were integrated as they synthesize complex physiological parameters into standardized indices widely used to predict ICU outcomes. The primary outcome was in-hospital mortality, defined as the patient’s survival status at the time of hospital discharge. The complete classification of extracted features is presented in Table 1. See supporting information S1 Table Comparison of features across prior studies for a comparison of feature sets across relevant studies and our initial candidate variables.

**Table 1.**
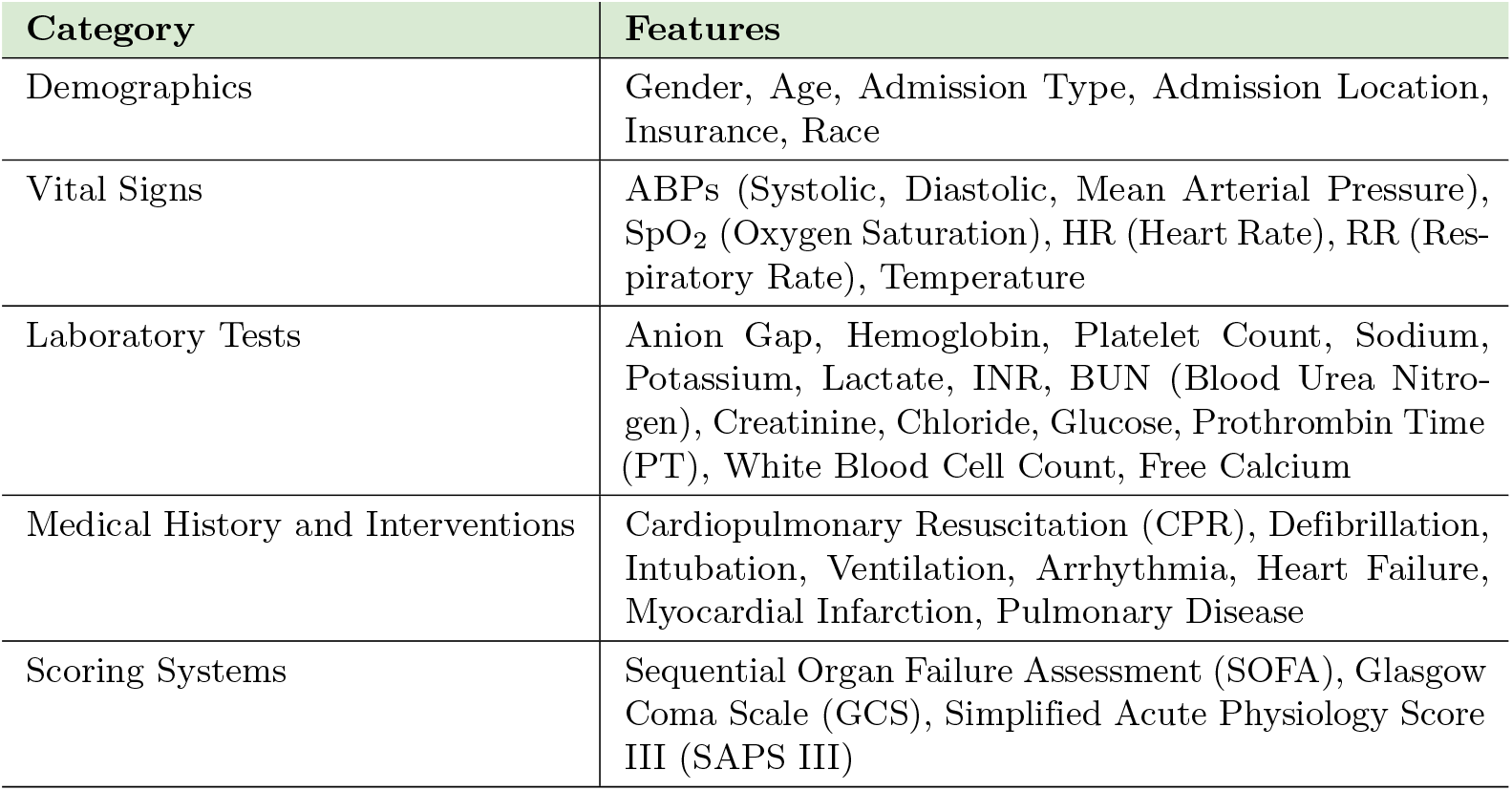
Extracted features for ICU mortality prediction.

Categorical variables were transformed using Label Encoding, a technique that converts categorical values into numerical representations suitable for machine learning models. Label Encoding assigns a unique integer to each category while preserving ordinal relationships when applicable. Our research utilized Label Encoding for healthcare datasets because it balances accuracy and computational cost well. It has advantages over One-Hot Encoding, another standard encoding method, which significantly increases feature dimensionality. In contrast, Label Encoding maintains a compact feature space, ensuring lower computational complexity while preserving model accuracy [36].

However, label encoding’s known limitation is its assumption of ordinal relationships, which may not exist in some categorical variables. This could potentially lead to misinterpretations in certain machine-learning models. To mitigate this issue, our research applied grouping techniques for categorical variables with a large number of unique categories. This approach reduced the total number of distinct values, minimizing bias and improving the model’s generalization while maintaining the benefits of a lower-dimensional representation.

To address missing data, a Random Forest-based imputation method was employed, which predicts missing values using patterns within the observed data. This approach ensures minimal data loss while effectively handling both categorical and numerical missing values. Unlike traditional imputation methods such as mean or mode imputation, Random Forest Imputation captures complex, nonlinear relationships between variables, preserving the variance in the dataset and reducing the risk of bias.

In epidemiological research, Random Forest-based imputation within the Multiple Imputation by Chained Equations (MICE) framework has demonstrated advantages over traditional parametric methods, particularly in datasets with nonlinearities and complex interactions [37]. For example, Pelgrims et al. [38] investigated discrepancies between self-reported and clinically measured health metrics and assessed the effectiveness of Random Forest-based multiple imputations in correcting biases in self-reported data when predicting hypertension and hypercholesterolemia. Their findings highlight the robustness of this method in reducing measurement errors and improving the accuracy of predictive models in health research.

### 2.4 Feature Selection

After extracting the features, feature importance selection was applied to identify the most clinically relevant predictors. Multiple complementary methods were employed, including LASSO regression, Ridge regression, ElasticNet, and Random Forest-based feature importance ranking, as illustrated in Figures 2. These techniques were selected to balance predictive performance with interpretability, which is essential in clinical applications where model transparency and trust are critical. LASSO regression was utilized for its ability to perform automatic feature selection by penalizing less informative variables, thereby reducing model complexity and highlighting key predictors with the strongest associations to mortality outcomes. Ridge regression was employed to stabilize coefficient estimates in the presence of multicollinearity, preserving clinically meaningful but correlated features that might otherwise be erroneously excluded. ElasticNet combined the strengths of LASSO and Ridge, effectively handling scenarios with multiple correlated clinical variables — a common characteristic of ICU datasets. Random Forest feature importance ranking provided a nonlinear perspective by capturing complex interactions among variables without assuming linearity, further enhancing the robustness of feature selection. By aggregating the results from these diverse methods, the final feature set was optimized to ensure statistical robustness, reduce overfitting risk, and maintain clinical interpretability, thereby supporting more accurate and actionable mortality predictions.

**Figure 2.**
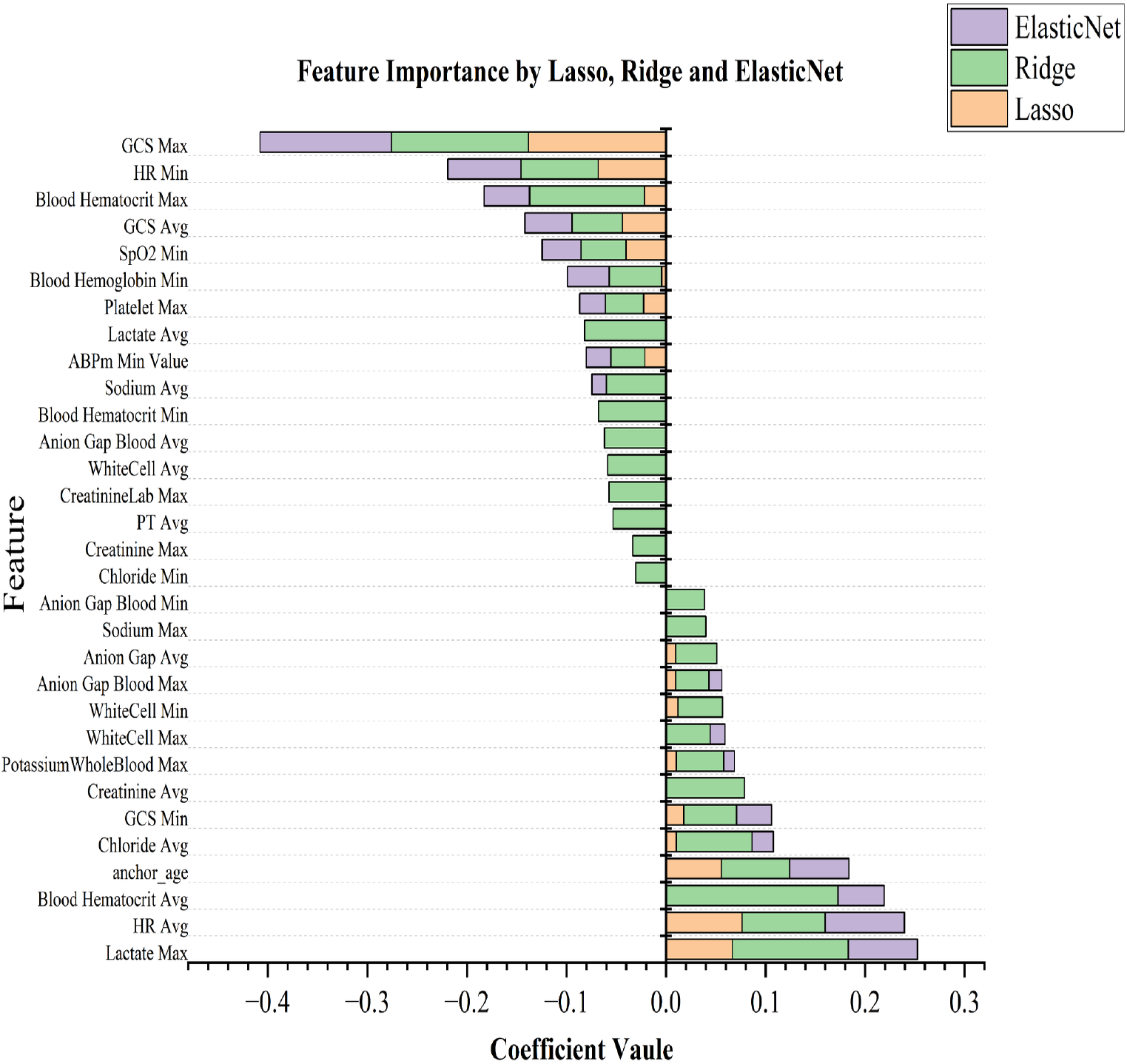
Feature importance ranking derived from LASSO regression, Ridge regression, and ElasticNet.

The final set of selected features was determined by combining the results of LASSO regression, Ridge regression, ElasticNet, and Random Forest Feature Importance (Importance *>* 0.01). To ensure optimal representation and avoid redundancy, only one version of each variable was retained. For instance, if both HR Min and HR Avg exceeded the selection threshold, only HR Avg was included in the final feature set.

This approach ensured that the most representative feature for each physiological parameter was used, enhancing model efficiency and interpretability.The final dataset comprised 2,385 patients with 17 features, selected through feature engineering and preprocessing as shown in Table 2.

**Table 2.** Extracted features for ICU mortality prediction.

| Category | Features |
| --- | --- |
| Demographics | Age |
| Vital Signs | SpO<sub>2</sub> Min, ABPm Min Value, HR Avg |
| Laboratory Tests | Lactate Max, Anion Gap Avg, Potassium (Whole Blood) Max, Creatinine Avg, Sodium Avg, Free Calcium Min, PT Avg, Blood Hemoglobin Min, Blood Hematocrit Max, INR Max, Chloride Avg, Platelet Max |
| Scoring Systems | GCS Max |

The baseline characteristics and extracted variables for all included patients are provided in Supporting Information S2 Table Summary of cohort characteristics at baseline.

## 3 Model Development and Statistical Analysis

### 3.1 Model Development

A range of supervised machine learning (ML) algorithms were developed and evaluated, including Logistic Regression, Extreme Gradient Boosting (XGBoost), Random Forest, Support Vector Machine (SVM), Adaptive Boosting (AdaBoost), Categorical Boosting (CatBoost), Bernoulli Naïve Bayes (NB), and K-Nearest Neighbors (KNN). Each model underwent hyperparameter tuning and optimization to enhance predictive performance.

To evaluate model efficacy, the discriminative performance of each model was quantified using the Area Under the Receiver Operating Characteristic Curve (AUC-ROC), which reflects the model’s ability to differentiate between survivors and non-survivors. In clinical practice, a high AUC-ROC is critical because it indicates reliable patient stratification, helping to prioritize those at highest risk for closer monitoring or early intervention. Additionally, our research assessed Precision, Recall, F1-score, and Accuracy to provide a comprehensive evaluation of classification performance across different thresholds. Precision is clinically important to minimize false positives, reducing unnecessary interventions that may expose patients to additional risks or resource burdens, while Recall ensures high detection of true high-risk cases, which is essential for avoiding missed opportunities for timely lifesaving care. F1-score balances these aspects, providing a harmonized view of clinical model reliability, and Accuracy offers an overall summary of correct predictions across the population. These complementary performance metrics ensured a balanced assessment of the models in terms of both sensitivity and specificity, both of which are crucial for clinical applicability, particularly in critical care environments where the consequences of false positives and false negatives are substantial.

Hyperparameter optimization was conducted via Grid Search with Cross-validation, focusing initially on the regularization parameter for the logistic regression model. Systematic hyperparameter tuning is essential for ensuring that models not only fit the training data but also generalize well to new, unseen patient populations. The model exhibited robust generalizability on the test dataset, achieving an AUROC exceeding 0.80, which indicates a high level of discriminative performance and supports its potential use in real-world clinical settings. Furthermore, the calibration curve was assessed to evaluate the reliability of probability estimates. Good calibration ensures that predicted mortality risks align closely with actual outcomes, which is critically important for clinical decision-making: interventions are often triggered based on specific risk thresholds, and over- or under-estimated risks could lead to inappropriate treatment strategies, patient harm, or resource misallocation.

### 3.2 Statistical Analysis

We introduced both the paired t-test and calibration assessment to evaluate the statistical robustness and clinical validity of our predictive models. These analyses ensure the models’ ability to generalize effectively to unseen patient populations, a critical factor for their clinical applicability and reliability. Specifically, the paired t-test was conducted to identify potential dataset shifts or biases by comparing the means of variables between the training and test datasets.

The paired t-test computes the test statistic as:

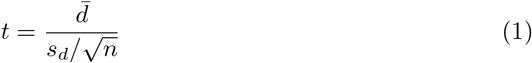

where 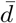 is the mean of the paired differences between training and test sets, *s*_*d*_ is the standard deviation of those differences, and *n* is the number of paired observations. A p-value less than 0.05 indicates a statistically significant difference between the paired means.

Out of the 17 analyzed variables, 14 variables showed no statistically significant differences between the datasets, suggesting consistent patient characteristics. However, 3 variables exhibited statistically significant differences, indicating the potential for dataset shift or bias that could influence model performance and generalizability.

Furthermore, the calibration analysis involved plotting calibration curves that depict the fraction of observed positive outcomes against predicted probabilities. Let 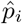 denote the predicted probability of positive outcome for the *i*-th patient, and *y*_*i*_ ∈ { 0, 1 } the actual outcome. The calibration function estimates:

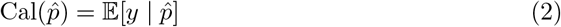

For a well-calibrated model, the calibration curve should approximate the identity line 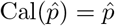. For the training dataset, the curve closely aligned with the diagonal reference, indicating strong calibration. However, the test dataset curve showed deviation in the mid-probability range, suggesting mild overconfidence in predicted risk.

These findings underscore the value of calibration assessments for identifying discrepancies between predicted and actual outcomes. Addressing such discrepancies is essential for improving model reliability in clinical decision-making and ensuring patient safety.

### 3.3 Feature Impact

To enhance the interpretability of the machine learning model developed for mortality prediction in CA patients, we introduced SHAP analysis. SHAP is a unified framework derived from cooperative game theory that provides locally accurate and consistent feature attributions for individual predictions. It decomposes the model output into additive contributions of each input feature, allowing us to quantify how much each variable influenced a specific prediction.

Formally, for a model *f* and input feature set *N*, the Shapley value for feature *i* is defined as:

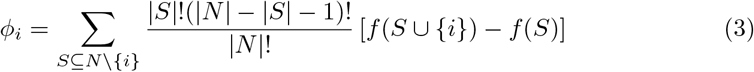

where *S* is a subset of the feature set not containing *i*, and *f* (*S*) represents the model prediction based only on features in *S*. The value *ϕ*_*i*_ represents the marginal contribution of feature *i* averaged over all possible feature combinations.

In clinical settings, interpretability of predictive models is particularly important, as it allows physicians to understand how and why the model arrives at certain predictions. This is critical for building trust, facilitating shared decision-making, and supporting risk-based patient stratification. Through SHAP analysis, we identified key predictive features—such as serum creatinine, blood pressure, and glucose variability—that significantly influenced model output. These insights not only improve transparency but also offer practical guidance for clinical interventions, potentially improving outcomes through more personalized and explainable decision support systems.

## 4 Results

### 4.1 Evaluation Results

Among the machine learning models assessed, CatBoost achieved the highest predictive accuracy, with an AUC-ROC of 0.9041 (95% CI: 0.8895–0.9188) on the training cohort and 0.8680 (95% CI: 0.8377–0.8926) on the test cohort. The model demonstrated a well-balanced trade-off between sensitivity (0.8479) and specificity (0.7021), reflecting strong discriminative capability in predicting in-hospital mortality.

To optimize the model, we conducted extensive hyperparameter tuning using GridSearchCV with 5-fold cross-test. The search space was carefully designed to enhance generalizability and mitigate overfitting. The following hyperparameter ranges were explored in Table 3:

**Table 3.** Hyperparameter ranges used for GridSearchCV optimization.

| Hyperparameter | Search Range |
| --- | --- |
| Iterations | 200, 300, 400 |
| Learning Rate | 0.02, 0.03 |
| Tree Depth | 2, 3 |
| L2 Regularization (l2_leaf_reg) | 10, 20, 30 |
| Bagging Temperature | 1, 5, 10 |
| Random Strength | 10, 20, 50 |
| Random Subspace Method (RSM) | 0.3, 0.5, 0.7 |

All models were trained on the same feature set derived from rigorous preprocessing and selection procedures. A stratified 70/30 train-test split was used to ensure balanced evaluation. The performance of each model was assessed using standard classification metrics such as AUC-ROC, accuracy, sensitivity, and specificity, the Predictive performance of all machine learning models show in the Table 4 The goal was to benchmark the relative predictive capabilities of these models and identify the optimal approach for clinical deployment in ICU mortality risk stratification.This comparative analysis enabled the identification of algorithms that not only demonstrated superior predictive accuracy but also maintained robust sensitivity–specificity balance.

**Table 4.** Predictive performance of machine learning models in the training and test sets.

| Model | AUC (95% CI) | Accuracy | Sensitivity | Specificity |
| --- | --- | --- | --- | --- |
| Training Set |  |  |  |  |
| Logistic Regression | 0.8728 (0.8555–0.8901) | 0.7975 | 0.8120 | 0.7766 |
| Random Forest | 0.8832 (0.8670 - 0.9003) | 0.7987 | 0.7602 | 0.8540 |
| XGBoost | 0.8709 (0.8532 - 0.8889) | 0.8083 | 0.8465 | 0.7533 |
| AdaBoost | 0.8874 (0.8711 - 0.9033) | 0.8191 | 0.8384 | 0.7912 |
| Bernoulli Naïve Bayes | 0.6152 (0.5904–0.6384) | 0.6285 | 0.7795 | 0.4117 |
| KNN | 0.8342 (0.8127–0.8520) | 0.7466 | 0.7612 | 0.7255 |
| SVM | 0.8802 (0.8641–0.8973) | 0.8071 | 0.8069 | 0.8073 |
| CatBoost | 0.9042 (0.8897–0.9190) | 0.8304 | 0.8567 | 0.7927 |
| Test Set |  |  |  |  |
| Logistic Regression | 0.8620 (0.8301 - 0.8880) | 0.7849 | 0.8272 | 0.7199 |
| Random Forest | 0.8513 (0.8194 - 0.8776) | 0.7905 | 0.7995 | 0.7766 |
| XGBoost | 0.8503 (0.8207 - 0.8760) | 0.7723 | 0.8525 | 0.6489 |
| AdaBoost | 0.8492 (0.8171 - 0.8753) | 0.7877 | 0.8410 | 0.7057 |
| Bernoulli Naïve Bayes | 0.6322 (0.5930–0.6673) | 0.6480 | 0.8226 | 0.3794 |
| KNN | 0.8377 (0.8061–0.8637) | 0.7584 | 0.7972 | 0.6986 |
| SVM | 0.8639 (0.8334–0.8897) | 0.7863 | 0.8134 | 0.7447 |
| CatBoost | 0.8680 (0.8377–0.8926) | 0.8120 | 0.8479 | 0.7021 |

The optimized CatBoost model employed 400 boosting iterations, a learning rate of 0.03, tree depth of 3, L2 regularization set to 10, a bagging temperature of 1, random strength of 20, and a random subspace method (RSM) value of 0.5. This configuration achieved the highest cross-validated AUC of 0.9041 on the training data and maintained robust generalizability with an AUC of 0.8660 on the test data. Collectively, these findings indicate that CatBoost offers superior performance for ICU mortality risk prediction, balancing discriminative power, generalization capability, and model interpretability.

### 4.2 Model Comparison

Conducted a comprehensive evaluation of five machine learning algorithms, each fine-tuned through grid search with cross-validation.

1. Logistic Regression (LR), a classic linear classifier widely used for binary outcomes, served as a strong baseline. The model with L2 regularization (`C=10, solver=‘liblinear’`) achieved an AUC of 0.8728 (95% CI: 0.8555–0.8901) on the training set and 0.8620 (95% CI: 0.8301–0.8880) on the testing set. It offered a balanced performance with good interpretability, making it suitable for clinical settings.
2. Random Forest (RF), an ensemble learning technique that constructs multiple decision trees and aggregates their outputs, was also evaluated. The optimal RF configuration (`n estimators=50, max depth=4`) yielded an AUC of 0.8832 (95% CI: 0.8670–0.9003) on the training set and 0.8513 (95% CI: 0.8194–0.8776) on the test set. The model exhibited high specificity, reflecting its robustness in correctly identifying patients who survived.
3. XGBoost is an advanced gradient boosting framework known for its computational efficiency and flexible regularization mechanisms. The optimal configuration (`max depth=3, learning rate=0.05, reg alpha=10, reg lambda=100`) achieved an AUC of 0.8709 (95% CI: 0.8532–0.8889) on the training dataset and 0.8503 (95% CI: 0.8207–0.8760) on the test dataset. These results highlight XGBoost’s competitive predictive performance, along with its capacity for effective overfitting control through regularization.
4. AdaBoost is an ensemble technique that sequentially adjusts sample weights to combine weak base learners—typically shallow decision trees—into a strong predictive model. The optimized configuration (`learning rate=0.02, n estimators=125`) achieved a training AUC of 0.8874 (95% CI: 0.8711–0.9033) and a test AUC of 0.8492 (95% CI: 0.8171–0.8753). Notably, the model demonstrated high sensitivity, indicating strong potential for reducing false-negative predictions in clinical risk assessment.
5. SVM with a linear kernel was optimized over C and gamma parameters. After incorporating standard scaling and extended grid search, it achieved a test AUC of 0.8462(95% CI: 0.8334 - 0.8897) and balanced sensitivity and specificity. Though slightly below ensemble models in performance, SVM maintained stability with fewer hyperparameters.

Overall, the comparative results (Figure 3)indicate that ensemble-based models—particularly CatBoost, Random Forest, and AdaBoost—consistently outperformed traditional classifiers in terms of both discrimination and calibration metrics. These methods demonstrated greater capacity to capture nonlinear relationships and complex interactions among ICU clinical variables. Logistic Regression and SVM, while offering advantages in interpretability and training efficiency, exhibited slightly lower AUCs and narrower specificity–sensitivity margins, suggesting limitations when modeling heterogeneous clinical outcomes.

**Figure 3.**
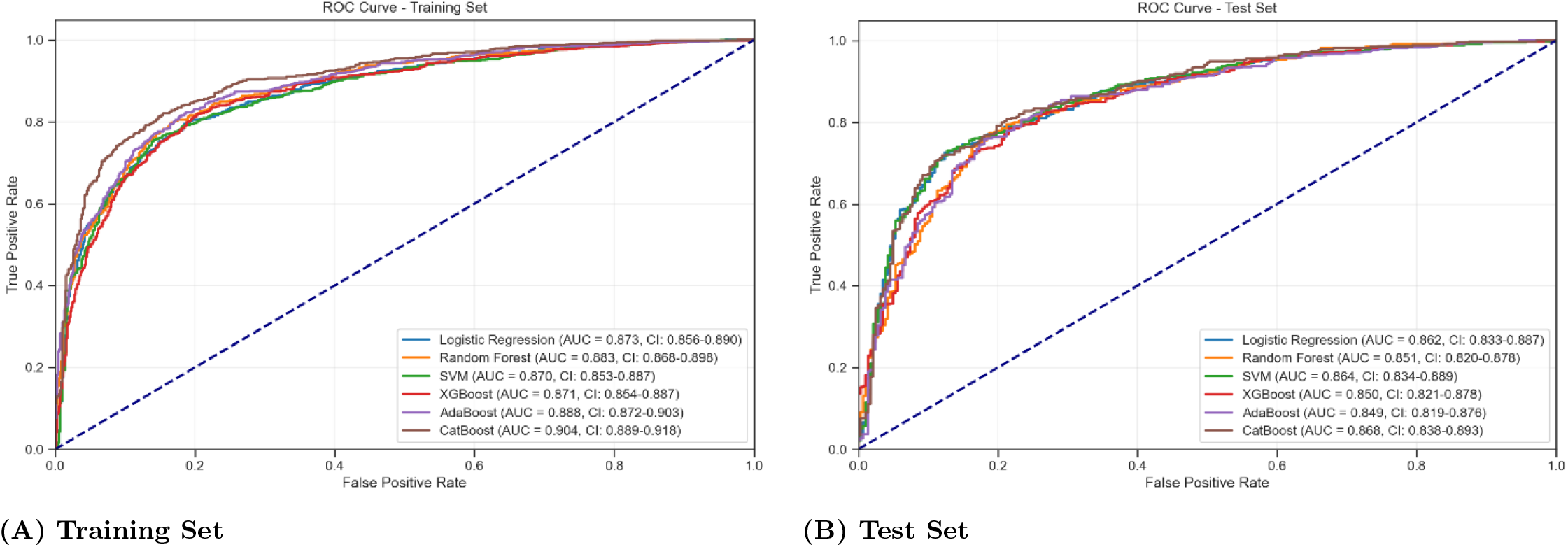
Model performance results across different algorithms on the training and test sets.

Importantly, the diversity in model architectures also contributed to complementary strengths. For instance, AdaBoost achieved high sensitivity, reducing the risk of false negatives, whereas Random Forest yielded superior specificity, beneficial for minimizing unnecessary interventions. The robustness and generalizability of these models, supported by cross-validation and hyperparameter tuning, provide a strong foundation for clinical deployment. These findings highlight the importance of algorithm selection in the development of reliable and interpretable ICU mortality prediction tools.

### 4.3 T-Test Results

We conducted independent two-sample t-tests for continuous variables. These tests aimed to identify any significant distributional shifts that could introduce sampling bias and adversely impact model performance, as shown in Table 5.

**Table 5.** T-test comparison of feature distributions between training and test sets.

| Feature | Train Mean | Test Mean | P-value |
| --- | --- | --- | --- |
| Lactate Max | 6.9874 | 7.7024 | 0.001667 |
| Anion Gap Avg | 15.7639 | 16.3715 | 0.007516 |
| Potassium WholeBlood Max | 5.0949 | 5.2334 | 0.012905 |
| GCS Max | 12.2475 | 11.8604 | 0.061530 |
| SpO<sub>2</sub> Min | 76.4693 | 74.9094 | 0.124051 |
| ABPm Min Value | 39.0839 | 37.8445 | 0.204271 |
| Creatinine Avg | 1.8389 | 1.8676 | 0.317509 |
| Sodium Avg | 138.8594 | 139.0189 | 0.398741 |
| Free Calcium Min | 1.0592 | 1.0632 | 0.424684 |
| PT Avg | 17.7216 | 17.5049 | 0.570737 |
| Blood Hemoglobin Min | 8.3431 | 8.2974 | 0.661737 |
| Blood Hematocrit Max | 41.1400 | 41.2467 | 0.704192 |
| HR Avg | 85.6622 | 85.4209 | 0.722243 |
| Anchor Age | 64.5842 | 64.7849 | 0.782111 |
| Platelet Max | 357.9066 | 356.7194 | 0.880769 |
| INR Max | 2.8217 | 2.8265 | 0.966567 |
| Chloride Avg | 102.5620 | 102.5662 | 0.984703 |

**Table 6.**
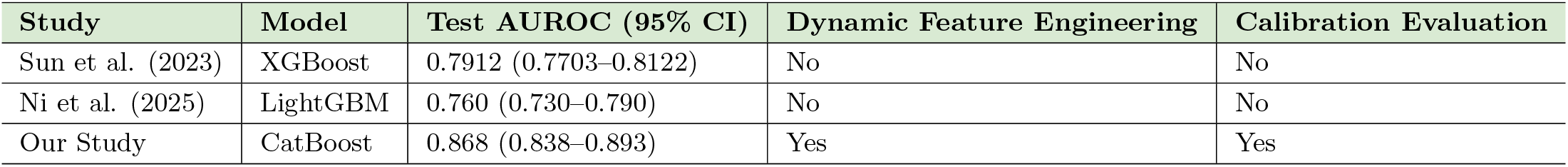
Comparison of model performance and methodology across studies.

For the continuous variables, t-tests were conducted to compare the means between the training and test sets. A significance threshold of p=0.05 was used to determine statistical significance. Among the 17 features analyzed, three exhibited statistically significant differences. This allowed for clearer comparison of relative differences across features without altering the statistical testing itself:

Lactate Max (p = 0.001667)

Anion Gap Avg (p = 0.007516)

Potassium (Whole Blood) Max (p = 0.012905)

Several potential factors may explain these discrepancies. For Lactate Max, temporal or cohort-based shifts in patient acuity may have influenced the distribution according to [39]. For instance, if the test set includes a higher proportion of patients admitted during periods of increased clinical severity (e.g., surges in septic shock or cardiac arrest), elevated lactate levels may result.

In the case of Anion Gap Avg, variations may be attributable to differences in the prevalence of renal dysfunction or other metabolic disorders between cohorts [40]. Additionally, changes in laboratory calibration protocols or updates to the electronic health record system may have introduced systematic biases in recorded values.

Finally, the observed difference in Potassium (Whole Blood) Max may reflect a greater incidence of renal impairment, diuretic use, or sample hemolysis in one of the datasets. Moreover, inconsistent utilization or availability of whole blood potassium measurements across time could also contribute to distributional shifts [41].

### 4.4 Calibration Test Results

To evaluate the reliability and probabilistic accuracy of the model’s predictions, calibration analyses were performed for both the training and testing datasets. Figure 4 illustrates the calibration curves, plotting the mean predicted probability against the observed fraction of positive outcomes across probability bins. The dashed diagonal line represents perfect calibration, where predicted probabilities would exactly match observed event rates.

**Figure 4.**
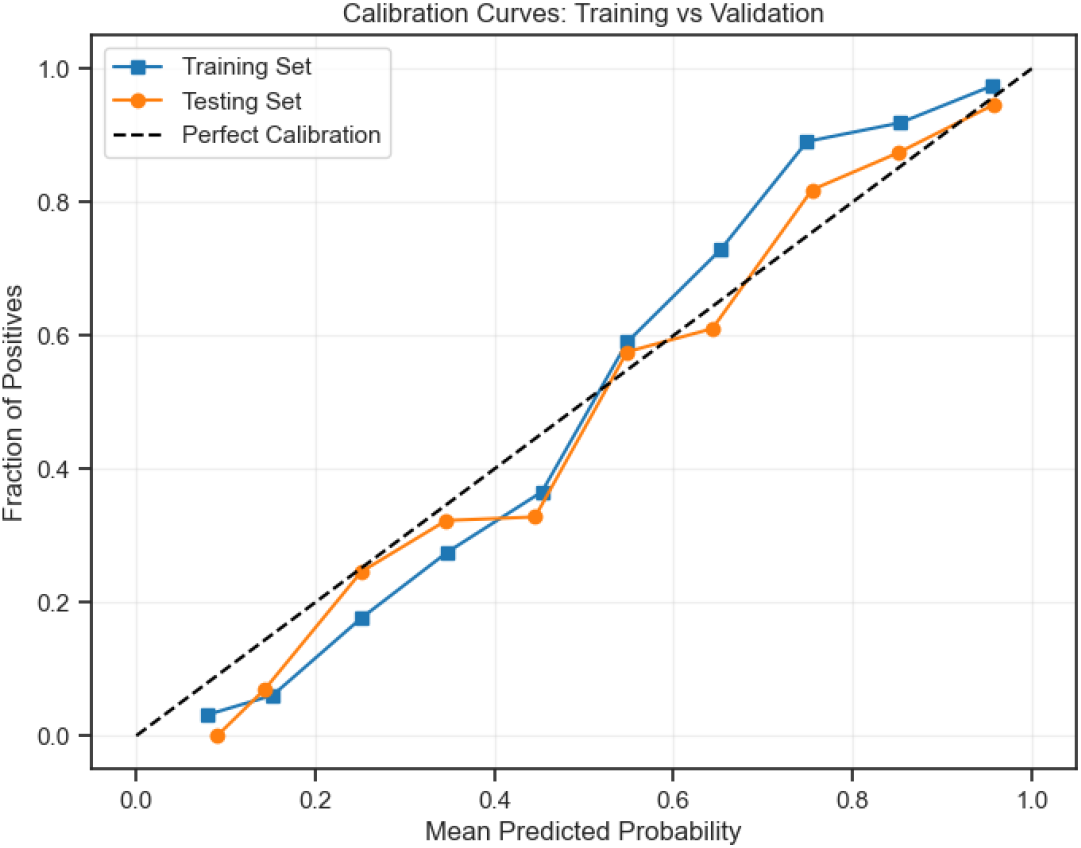
Calibration curve for model evaluation on the test set.

In the training cohort, the calibration curve closely follows the ideal diagonal across most probability ranges, with slight deviations observed in the lower predicted probability bins (0–0.2) where mild underestimation occurred, and at the higher end (above 0.8) where minor overconfidence was noted. In the testing cohort, the calibration curve similarly approximates the perfect calibration line, albeit with slightly greater variability, particularly in the mid-range (0.4–0.6) predicted probabilities. Nonetheless, the overall alignment between predicted and observed probabilities in both cohorts suggests satisfactory model calibration.

Quantitative assessment using the Brier score—a proper scoring rule measuring the mean squared error between predicted probabilities and actual outcomes—further corroborated these findings. The model achieved a Brier score of 0.127 on the training set and 0.143 on the testing set, indicating good overall calibration and minimal overfitting. The marginally higher Brier score in the testing set reflects expected slight degradation in calibration when applied to unseen data but remains well within acceptable bounds for clinical risk prediction models.

These calibration results highlight the model’s robustness in probabilistic forecasting, ensuring that predicted risks are meaningfully interpretable by clinicians. However, they also emphasize the necessity for continuous monitoring and recalibration if the model is deployed across different hospital settings or patient populations, where covariate shifts may compromise calibration integrity. Such ongoing assessment would be crucial for maintaining predictive reliability and maximizing clinical utility in real-world environments.

### 4.5 Shape Analysis

A SHAP analysis was performed on the final model to enhance understanding of the model’s decision-making process and feature influence. SHAP values provide a unified framework for interpreting individual feature contributions to model predictions, enabling both global and instance-level interpretability, as illustrated in Figures 5. This is particularly important in high-stakes clinical settings where model transparency can enhance clinician trust and facilitate clinical decision-making.

**Figure 5.**
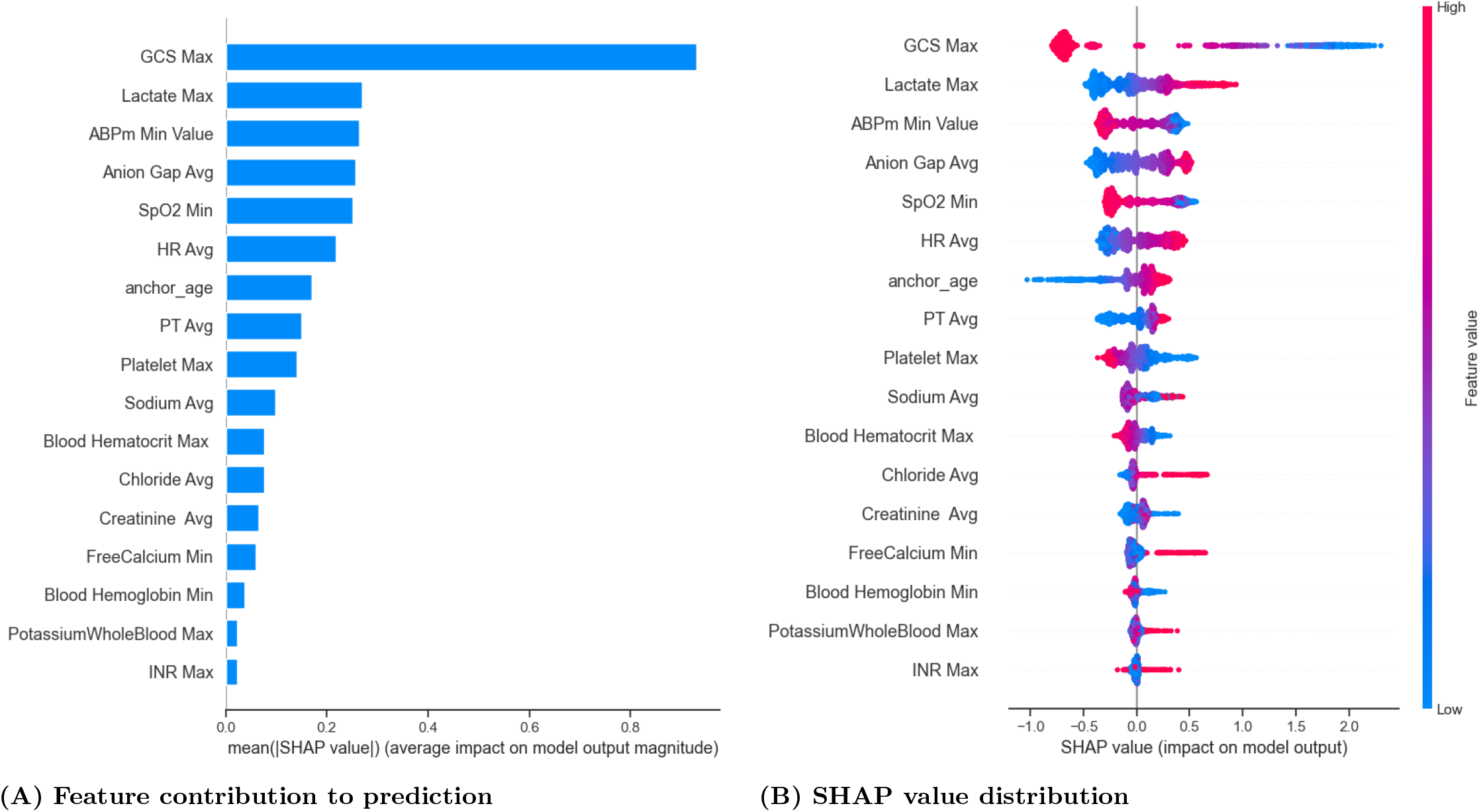
SHAP-based interpretation of model predictions.

The SHAP summary bar plot highlights the most influential predictors of mortality, ranked by their mean absolute SHAP value (i.e., average impact on model output magnitude). The most important features were:

GCS Max (Glasgow Coma Scale): the strongest predictor, with lower values associated with higher mortality risk.

Lactate Max: elevated lactate levels contributed positively to predicted mortality, reflecting metabolic stress or hypoperfusion.

ABPm Min Value (Arterial Blood Pressure Minimum): lower values increased risk, consistent with hemodynamic compromise.

Anion Gap Avg and SpO_2_ Min also had considerable influence, suggesting that metabolic acidosis and hypoxia were key mortality indicators.

The beeswarm plot further illustrates how feature values influence predictions. For instance, high values of GCS Max (blue) were associated with negative SHAP values, reducing the predicted risk, while low GCS values (pink) substantially increased predicted mortality. Similarly, higher Lactate Max and Anion Gap Avg values were associated with positive SHAP values, pushing predictions toward higher risk.

## 5 Discussion

### 5.1 Summary of existing model compilation

Accurate prediction of in-hospital mortality following cardiac arrest is crucial for improving ICU patient management and optimizing resource allocation. This study developed ML pipelines using the MIMIC-IV database to forecast in-hospital death among ICU patients after cardiac arrest. Our work provides both methodological advancements and clinical implications by integrating rigorous feature selection, robust modeling, model calibration, and interpretability analyses.

Compared to traditional methods like the Modified Early Warning Score (MEWS) and logistic regression-based scores (e.g., APACHE II), our model captures nonlinear relationships and dynamic feature patterns, resulting in higher discriminative performance (test AUC 0.8680 vs. traditional AUCs 0.7). Additionally, while conventional methods use a static feature snapshot, our model incorporates maximum, minimum, and average trends during ICU stay, a clinically valuable enhancement for dynamic risk stratification.

Previous predictive models for post-cardiac arrest outcomes often suffered from small sample sizes, lack of external validation, or limited feature sets that ignored dynamic physiological changes. By contrast, our study leveraged a large, contemporary ICU cohort, systematically extracted comprehensive clinical features, and applied sophisticated feature engineering techniques. Statistical aggregations (mean, max, min) were utilized to capture the temporal variability of physiological parameters, an approach clinically valuable for modeling the dynamic instability of ICU patients. Label encoding and Random Forest-based imputation ensured that both categorical and missing data were handled with high fidelity, minimizing bias and maximizing clinical representativeness.

Feature selection was performed using a combination of LASSO, Ridge, ElasticNet regularization, and Random Forest importance scores. This multi-method feature selection strategy enhanced robustness, reduced overfitting risk, and prioritized clinically meaningful variables such as GCS, lactate, and blood pressure parameters, which are known critical markers of ICU mortality risk. Clinically, narrowing down to the most predictive features facilitates interpretability and practical deployment of predictive tools in real-time ICU settings.

Eight supervised ML algorithms were developed and compared. CatBoost emerged as the best-performing model, achieving a training AUC of 0.9041 and a test AUC of 0.8680, outperforming traditional models like logistic regression. Importantly, model calibration was assessed, revealing strong probability alignment on the training set and minor overconfidence on the test set, suggesting high clinical reliability but also underscoring the need for recalibration when deployed across different cohorts.

SHAP analysis further improved model transparency, revealing that neurological status (GCS), metabolic parameters (lactate, anion gap), and hemodynamic instability (ABP minimum) were the dominant mortality predictors. These findings align with clinical knowledge, thus reinforcing physician trust in the model and enhancing its potential as a clinical decision support tool. Through SHAP, physicians can understand individualized patient risks rather than relying on black-box outputs, thereby supporting shared decision-making and patient-specific interventions.

Our statistical tests confirmed the absence of major distribution shifts between training and test cohorts for most features, enhancing model generalizability. However, minor shifts in lactate and potassium levels were noted, highlighting the necessity for ongoing calibration checks when applying models across temporal or institutional settings.

### 5.2 Comparison with previous studies

Recent studies have attempted to predict in-hospital mortality among ICU patients after cardiac arrest using machine learning methods. Sun *et al*. [42] constructed models based on the MIMIC-IV database, achieving a test AUROC of 0.7912 (95% CI, 0.7703–0.8122). Ni *et al*. [43] reported a test AUROC of 0.760 (95% CI, 0.730–0.790) using a LightGBM model. However, both studies primarily relied on static admission features, and neither incorporated advanced dynamic feature engineering, calibration evaluation, or systematic explainability analyses, potentially limiting their clinical applicability.

By contrast, our study significantly outperformed these prior works. Our CatBoost model achieved a test AUROC of 0.868 (95% CI, 0.838–0.893) and a Brier score of 0.143 (95% CI, 0.134–0.152), indicating not only superior discrimination but also better probability calibration. Furthermore, our model achieved a test accuracy of 0.812, substantially higher than the accuracies reported in Sun *et al*. and Ni *et al*., reflecting improved consistency in patient outcome prediction.

Importantly, beyond numerical performance improvements, our approach introduced dynamic feature aggregation (capturing maximum, minimum, and average trends), rigorous feature selection using RFECV and LASSO, and full SHAP-based explainability, providing individualized risk interpretations. These methodological innovations address key gaps in previous studies and enhance the clinical utility of our model for ICU decision support.

### 5.3 Limitations and Future Work

Despite these strengths, our study has limitations. Although the model demonstrates strong internal validation within MIMIC-IV, external validation across multiple centers is essential before clinical adoption. Additionally, while we captured time-aggregated statistics, richer time-series modeling (e.g., using RNNs) may further enhance predictive performance by fully exploiting temporal trends in ICU data.

Future work should focus on external validation, prospective evaluation in real-time ICU workflows, and development of dynamic risk prediction models incorporating continuous physiological monitoring. Furthermore, integrating imaging or waveform data could uncover additional latent predictors and enrich model performance.

## 6 Conclusion

This study systematically addressed the challenge of predicting in-hospital mortality in ICU patients after cardiac arrest by implementing a comprehensive and interpretable machine learning pipeline. Researchers utilized the MIMIC-IV database to construct a retrospective cohort of 2,385 patients, extracting dynamic clinical features through statistical summarization of ICU stay data. Data preprocessing included Random Forest-based imputation for missing values and structured encoding for categorical variables. A rigorous feature selection process combining LASSO, Ridge, ElasticNet, and Random Forest importance ranking refined the feature set to 17 clinically relevant variables, including Glasgow Coma Scale (GCS), serum lactate, and arterial blood pressure minimum.

To enhance model robustness and fairness, stratified 70/30 train-test splits were employed, with hyperparameter tuning conducted via GridSearchCV. Eight supervised machine learning models were benchmarked, and CatBoost achieved superior performance, with an AUC of 0.904 on the training set and 0.868 on the test set, outperforming traditional classifiers like Logistic Regression, Random Forest, and XGBoost. Calibration curve assessments and paired t-tests ensured statistical consistency and minimized dataset shift. Additionally, SHAP analysis provided transparent interpretation of model predictions, identifying GCS, lactate, and anion gap as key mortality predictors.

These findings demonstrate that machine learning, when carefully constructed with interpretable features and rigorous validation, can effectively augment clinical decision-making in critical care settings. Future research should focus on external validation across diverse populations and real-time deployment to enhance the model’s generalizability and clinical integration.

## Supporting information

**S1 Table Comparison of features across prior studies**. This table compares the clinical features reported in recent studies on ICU mortality prediction with those used in our study…

**S2 Table Summary of cohort characteristics at baseline**. This table presents demographic, vital signs, laboratory, and comorbidity information for patients included in the final analytic cohort. Features were summarized using mean (±SD) for continuous variables and count (percentage) for categorical variables.

## Data Availability

The data used in this study were obtained from the publicly available MIMIC-IV database (https://physionet.org Accurate ide), which requires credentialed access. No new data were generated.

## Ethics Statement

This study was conducted using the MIMIC-IV database, a de-identified and publicly available dataset. Ethical approval was not required. Access was granted under the credentialed data use agreement (Certification Number: 50778029). Patient consent was waived as the dataset used is fully de-identified and publicly available.

